# Quantitative Dixon-Based PDFF and R2* Estimation and Optimization on MR-Simulation and MR-Linac Devices for the Pelvis and Head and Neck: A Prospective R-IDEAL Stage 0-2a Study

**DOI:** 10.64898/2026.03.09.26347965

**Authors:** Lucas McCullum, Natalie A. West, Kayeong Shin, Brian A. Taylor, Alexander Augustyn, Omran Saifi, Sara Thrower, Jihong Wang, Shalin Shah, Seungtaek Choi, Chidinma P. Anakwenze, Clifton D. Fuller, Warren Floyd

**Author notes:** Corresponding Author: Warren Floyd.

## Abstract

**Background and Purpose:** The use of MRI-based fat quantification can be applied to automatically identify red bone marrow which is highly sensitive to radiation and systemic therapies and could be used as an organ-of-interest for adaptive radiation therapy. Currently, the tradeoff of scan time and PDFF/R2* quantification accuracy from the 2-/3-/6-point methods, particularly for the time-constrained MR-Linac, remain unanswered. Therefore, the purpose of this study was to investigate the technical feasibility and quantitative performance of quantitative Dixon-based imaging for scanners within the radiation oncology department.

**Materials and Methods:** A 2-/3-/6-point version of the quantitative Dixon sequence was developed and scanned on a 1.5T MR-Simulation, 3T MR-Simulation, and 1.5T MR-Linac scanner for five repetitions using the Calimetrix Model 725 PDFF-R2* phantom as a nominal reference for quantitative PDFF/R2* values. The image geometric distortion as well as the quantitative concordance, Bland-Altman agreement, repeatability, and reproducibility of both the PDFF/R2* values were determined. Each sequence was evaluated in both the pelvis and head and neck across both healthy volunteers and patients.

**Results:** The most severe geometric distortion was less than 2 mm except for the 1.5T MR-Linac when using the 2-point Dixon sequence with distortions exceeding 5 mm. The 6-point Dixon sequence showed the highest concordance at above 0.97 across all scanners for both PDFF and R2* followed by the 3-point and 2-point sequence. The 2-point Dixon sequence exhibited significant PDFF biases particularly at the higher R2* values since it did not correct for it during reconstruction. For the Bland-Altman analysis, the 2-point Dixon sequence had the widest 95% limits of agreement followed by the 3-point and 6-point Dixon sequence with the narrowest bands. The goodness-of-fit is generally lowest at higher PDFF values and lower R2* values. Both repeatability and reproducibility were the lowest for the 6-point Dixon sequence.

**Discussion:** The 6-point quantitative Dixon sequence demonstrated superiority for the chosen evaluation metrics. The results of this work can be used to determine the threshold for true quantitative changes of PDFF/R2* while considering acquisition variabilities, enabling future biomarker studies and clinical trials. Further, this work provides validation for future investigations into quantitative bone marrow characterization.

**Graphical Abstract:** 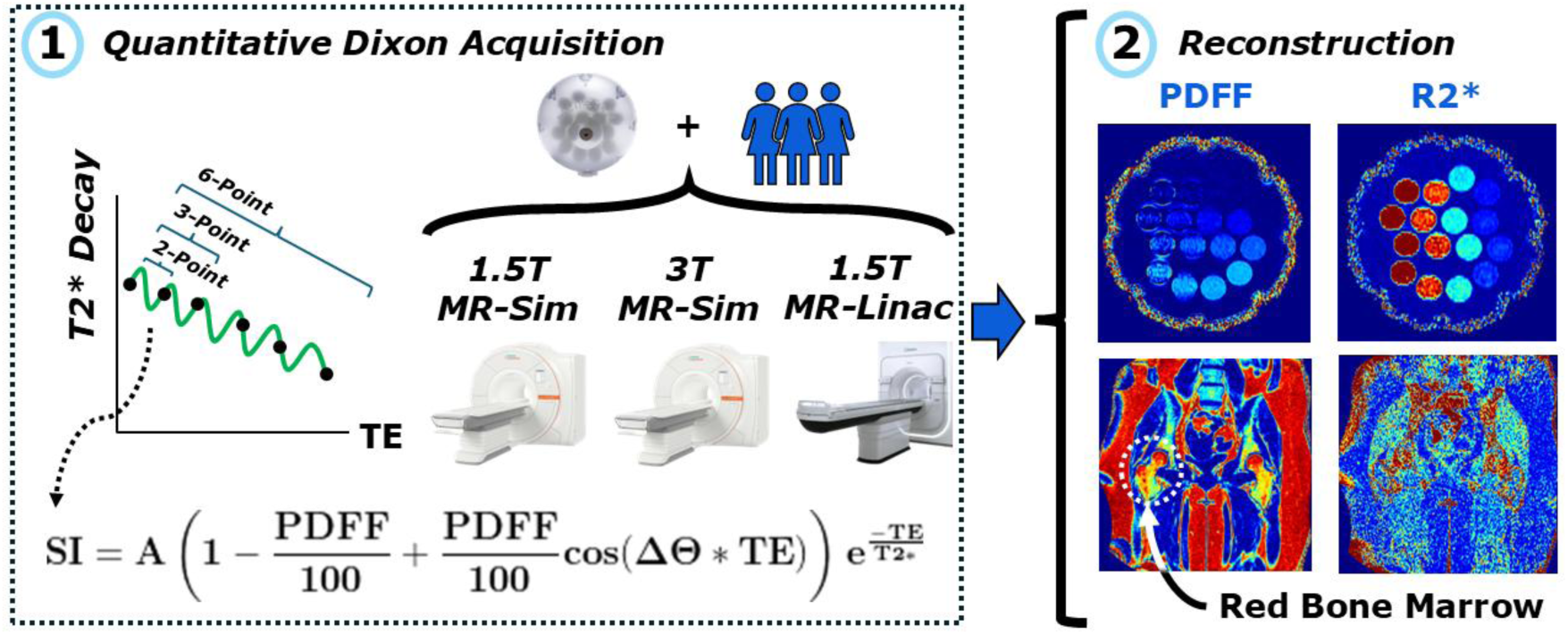

## 1. Introduction

Fat is present nearly everywhere throughout the body in varied compositions^1^ leading to many questions regarding its changes in response to radiation therapy and how this can be used to adapt radiation therapy treatment plans^2,3^. The most preferred technique for fat quantification is magnetic resonance imaging (MRI) due to its reduced ionizing radiation exposure compared to computed tomography (CT), especially with serial measurements^4,5^. These serial measurements are necessary in radiation oncology to monitor radiation-induced side effects and derive longitudinal imaging biomarkers for treatment adaptation^6–18^. They can be performed using a combination of an MRI-based simulation (MR-sim) device for initial treatment planning and an integrated MRI and linear accelerator (MR-Linac) device for combined imaging and radiation therapy treatment delivery^19,20^. The benefit of this combination compared to traditional CT-based simulation is the allowance for an MR-only workflow which provides better tumor conspicuity while enabling longitudinal tracking on top of the reduced ionizing radiation exposure^21–26^.

Proton density fat fraction (PDFF) mapping in MRI is typically done by exploiting the inherent resonant frequency difference between fat and water, the two most common MRI-visible substances in the body, in a technique known as the Dixon method^27^. This resonant frequency difference means that, within each voxel, there is a time that the fat and water spins align and both contribute to the signal (in-phase) and another time where their spins are opposite and reduce the overall signal (opposed-phase). These two images can then be added together to create a water-only image or subtracted from each other to generate a fat-only image in a technique known as the 2-point Dixon method due to its use of two images along the fat and water dephasing cycle. The fastest way to do this on modern MRI systems is through gradient echo imaging. Unfortunately, many assumptions are used for the 2-point Dixon method, so its use for accurate quantitative fat determination is limited by static magnetic field inhomogeneity which will affect the observed resonant frequencies, R2* decay from gradient echo free induction decay (FID) which will affect signal intensity, and the introduction of T1 signal from the flip angle used during readout which will also affect the signal intensity^28,29^. Due to these limitations, 3-point Dixon methods have been proposed which can simultaneously quantify R2* decay and static magnetic field inhomogeneity while correcting the fat fraction^30,31^. This has been expanded to a 6-point Dixon method which can better identify the multiple fat resonant peaks^32^, thus providing a more accurate representation of fat within the body instead of just the bulk methylene peak which is most prominent and the basis for the 2-point Dixon method^33^. However, a critical question remains in its application to radiation oncology with the tradeoff of more accurate R2* and PDFF quantification from the 3-point and 6-point methods against their longer scan times, particularly for the time-constrained MR-Linac.

The applications of MRI-based fat quantification to the field of radiation oncology are wide. One growing application is the automated identification and monitoring of sarcopenia, or the progressive loss of muscle mass^34^, which has been shown to correlate with inferior outcomes in radiation therapy^35^. This field has grown rapidly with more recent automated methods segmenting internal structures necessary for its calculation from MRI images^36–40^. Another potential application of fat quantification from MRI is in the quantitative assessment of bone marrow^41,42^, which is affected by both radiation^43,44^ and systemic therapies^45^, and could be used as an organ-of-interest for adaptive radiation therapy. Specifically, differentiating between yellow (“inactive”) and red (“active”) bone marrow can be done with fat quantification techniques due to the difference in fat content which is ∼95% and ∼40%, respectively^46^. In place of fat, the red bone marrow contains hematopoietic cells and lymphocytes which are highly radiosensitive and critical to improve outcomes for patients, particularly in the case of lymphopenia^47,48^. This concern has motivated several prospective studies evaluating both the technical feasibility and clinical outcomes of active bone marrow sparing with quantitative MRI as an ideal tool for its identification^49,50^. Further studies found that dose-volume metric to the active bone marrow is associated with decreased nadir of white blood cell counts and absolute neutrophil counts^51^.

This has been taken further by developing normal tissue complication probability (NTCP) models^52^ relating radiation dose to active bone marrow and direct outcomes such as nadir of white blood cell counts and absolute neutrophil counts^53,54^.

Several studies have validated (i.e., tested accuracy, repeatability, and reproducibility) of quantitative MRI-based Dixon acquisitions in phantoms for a wide range of diagnostic scanners^55–57^, however its translation to radiation oncology MR-simulation scanners is minimal^58,59^ with no studies identified by the authors for its clinical application on the MR-Linac except for 2-point techniques^60^ which is used primarily for synthetic-CT generation^61,62^.

Therefore, the purpose of this study is to investigate the technical feasibility of utilizing quantitative Dixon-based imaging as an R-IDEAL^63^ Stage 0-2a study. This will be achieved and shown by (1) validating R2* in phantoms, (2) validating PDFF in phantoms, and (3) comparing these performance metrics across 2-echo, 3-echo, and 6-echo acquisitions in both phantoms and healthy human subjects using both a 1.5T and 3T MR-sim scanner and a 1.5T MR-Linac scanner. The results of this work can be used to determine the threshold for true quantitative changes of both R2* and PDFF while considering acquisition variabilities, enabling future biomarker studies and clinical trials.

## 2. Methods and Materials

### 2.1. MRI Acquisition Parameters

All MRI scans were performed on the following scanners: 1) 1.5T Siemens MAGNETOM Sola Fit MR-Sim scanner (XA51; Siemens Healthcare; Erlangen, Germany), 2) 3T Siemens MAGNETOM Vida MR-Sim scanner (XA60; Siemens Healthcare; Erlangen, Germany), and 3) 1.5T Elekta Unity MR-Linac (R5.8.1; Elekta AB; Stockholm, Sweden). Each quantitative Dixon sequence utilized successive gradient / fast field echoes (GRE on Siemens / FFE on Philips) with the choice of echo times determined by optimizing for the number of signal averages^64^ (NSA) across the multiple echo times where {0, 2π, …} is in-phase and {π, 3π, …} is opposed-phase^28^ while maintaining achievability on our MRI scanners. Specifically, the 2-point technique^65,66^ would ideally use intervals of {0, π}, the 3-point technique^67,68^ would use intervals of {0, π/3, 2π/3}, and the 6-point technique^33^ would use intervals of {0, π/6, π/3, π/2, 2π/3, 5π/6} while maintaining the lowest possible echo times for the most accurate R2* quantification. A low flip angle of 3° was used to minimize the effects of T1 relaxation which can overestimate the PDFF values^29^. No image acceleration was used to minimize the effects of noise amplification^69^, which is also in line with how we scan clinical patients. The images were acquired in the coronal orientation. All other acquisition parameters were kept as similar as possible across the systems to enhance reproducibility. On both the 1.5T and 3T MR-Sim scanners, two homogeneous “Solution N” (3.75g NiSO_4_ x 6H_2_O and 5g NaCl per 1,000g H_2_O) phantoms had to be added to the superior and inferior area next to the phantom to prevent fat / water swaps due to the large field-of-view to accommodate the pelvis. In addition to the scanner reconstructed PDFF and R2* maps, for the MR-Sim scanners, a goodness-of-fit map was also reconstructed from the scanner to evaluate the residual fitting errors for the PDFF and R2* maps where smaller values are more reliable. Further acquisition details for both the proposed quantitative Dixon acquisitions can be seen in **Table 1**.

**Table 1.**
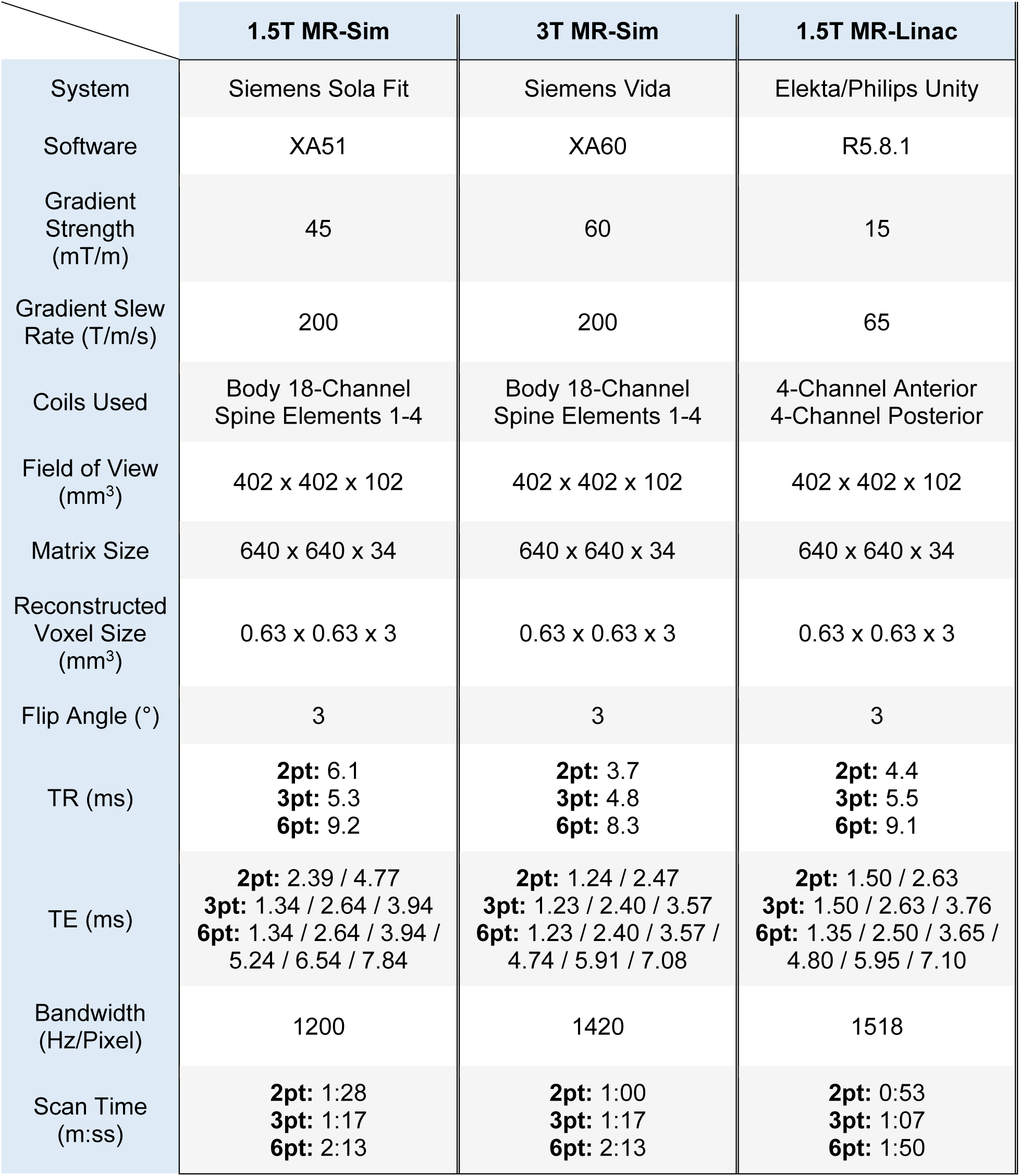
The MRI acquisition parameters for the proposed quantitative Dixon sequences. Abbreviations: pt = point, TE = echo time, TR = repetition time.

### 2.2. Phantom / Human Subject Assessment

We utilized the Calimetrix Model 725 PDFF-R2* phantom^70^ (Calimetrix; Madison, WI, USA) as a nominal reference for quantitative R2* and PDFF values. To ensure compliance with phantom nominal reference values, the temperature was monitored using an analog thermometer placed near the bore of the MRI which regularly read near 20°C across all acquisitions for each scanner. Repeatability and reproducibility in the Calimetrix phantom were conducted by scanning them separately for five consecutive repetitions across each MRI scanner. To evaluate geometric distortion, a critical factor when applying the maps for radiation therapy treatment planning, we measured the width and height of each vial inside the Calimetrix phantom using the PDFF map in ImageJ^71^ (1.54d; National Institutes of Health, MD, USA).

The optimal image quality clinically was determined by scanning two healthy human subjects and five patients (three pelvis, two head and neck) using the imaging protocol in **Table 1**. The participants provided written informed consent to an internal volunteer imaging protocol (PA15-0418), both approved by the institutional review board at The University of Texas MD Anderson Cancer Center. In the pelvis, the 2-/3-/6-point Dixon sequences were compared in a healthy volunteer and the iliacus muscle, subcutaneous adipose tissue, yellow bone marrow, and red bone marrow were chosen for analysis. Each structure was identified and a circular region-of-interest (ROI) was drawn using the respective PDFF or R2* maps in ImageJ.

### 2.3. Statistical Analysis

All relevant statistical analysis was computed and communicated while following the Statistical Analyses and Methods in the Published Literature (SAMPL) guidelines^72^. The R2* maps were determined automatically from the scanner for the 3-point and 6-point sequences while the PDFF maps were determined for all three echo time strategies. To evaluate the concordance between the quantitative Dixon sequences and the nominal phantom values, Lin’s Concordance Correlation Coefficient^73^ (LCCC) was computed for both the R2* and PDFF values. Bland-Altman analysis^74^ was conducted to assess limits of agreement. Repeatability and reproducibility was computed using the percentage ratio of the standard deviation to the mean, or the coefficient of variation (CoV), across the intra-/inter-scanner acquisitions, respectively. A graphical abstract of the proposed study is provided in **Figure 1**.

**Figure 1.**
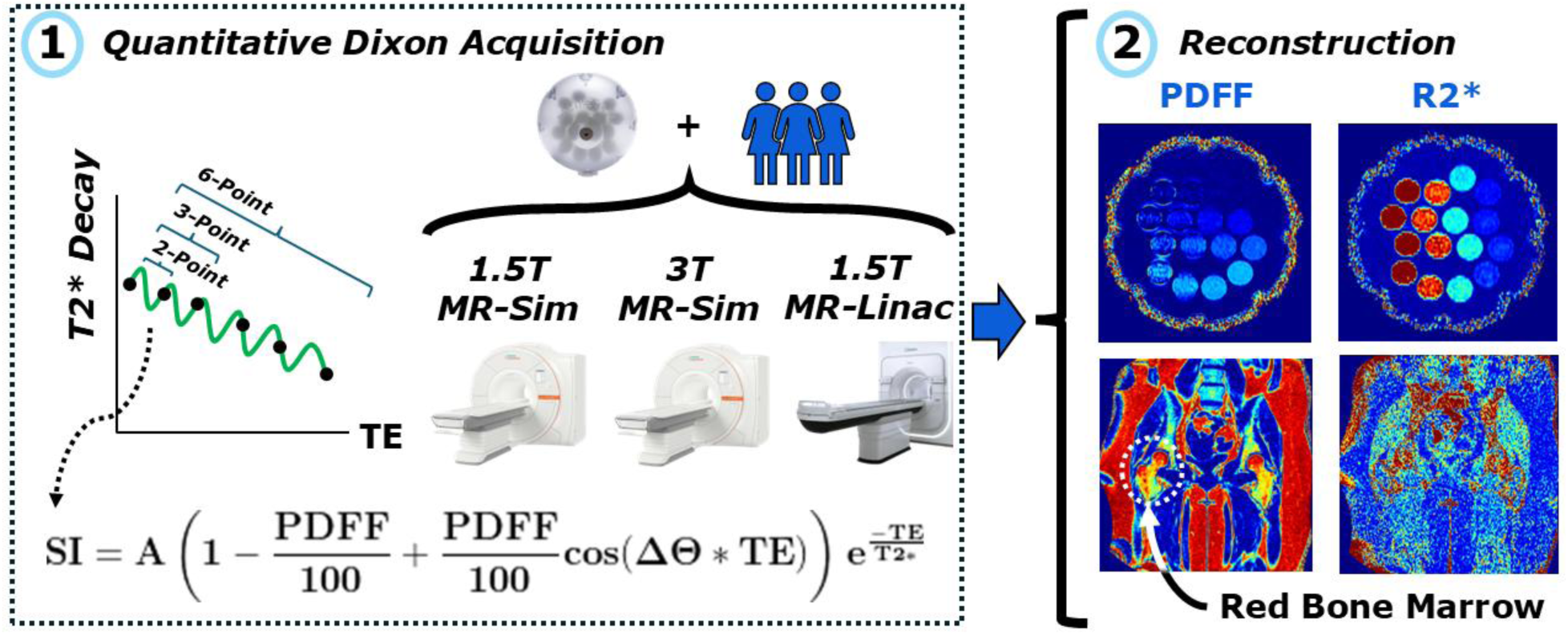
A graphical abstract of the proposed study where SI = signal intensity, A = initial signal amplitude, PDFF = proton density fat fraction, Ɵ = dephasing angle, and TE = echo time. (1) A series of 2-/3-/6-point Dixon sequences were acquired on a 1.5T MR-Sim, 3T MR-Sim, and 1.5T MR-Linac in both phantoms and human subjects. (2) Quantitative PDFF and R2* maps are reconstructed and assessed in phantoms and human subjects. Shown here is an example of red bone marrow determination in the pelvis. A specific example in the head and neck is not shown; however, it was included in the assessment for this study.

## 3. Results

### 3.1. Acquisition Parameter Results

The achieved dephasing angles superimposed on the theoretical optimal values for NSA are shown in **Figure 2**. Each value is near the optimal dephasing angle in terms of NSA with the slight exception of the 1.5T MR-Linac which is slightly below at approximately 1.5 NSA compared to the optimal 2. Similarly, the 3T MR-Sim was also difficult to achieve similar dephasing angles as on the 1.5T scanner due to the doubled dephasing resonance frequency.

**Figure 2.**
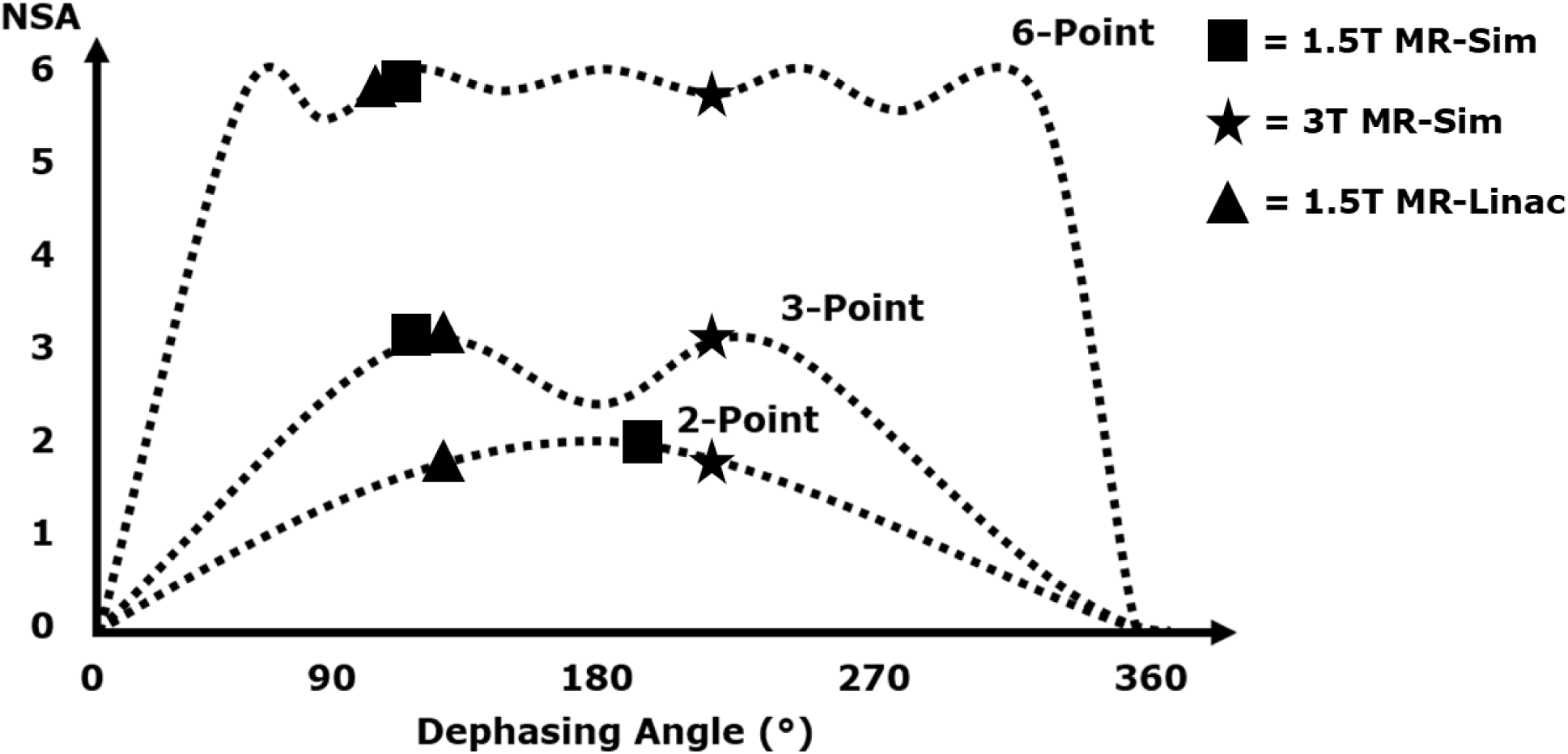
A demonstration of the achieved dephasing angles superimposed on the theoretical optimal values for NSA. The ideal values are the furthest peaks to the left due to their maximum NSA (sum of the number of echoes) as well as their fastest sampling of the T2* decay curve.

### 3.2. Phantom Results

The measured geometric distortion represented as the difference between the measured width and height of each vial in the Calimetrix phantom and the nominal diameter of 25 mm is shown in **Figure 3**. For most scanners and specific Dixon sequences tested, the calculated difference between the measured and nominal vial diameter was less than 2 mm. Some outlier values were noticed; however, the 1.5T MR-Linac had several measurements with a difference of 5 mm or more in the height direction. Upon evaluation of the images, these vials were normal in the width direction while being compressed in the height direction. Some heteroskedasticity was seen across the R2* values in the 3T MR-Sim scanner with increasing geometric distortion at the higher R2* values due to the induced magnetic field inhomogeneities.

**Figure 3.**
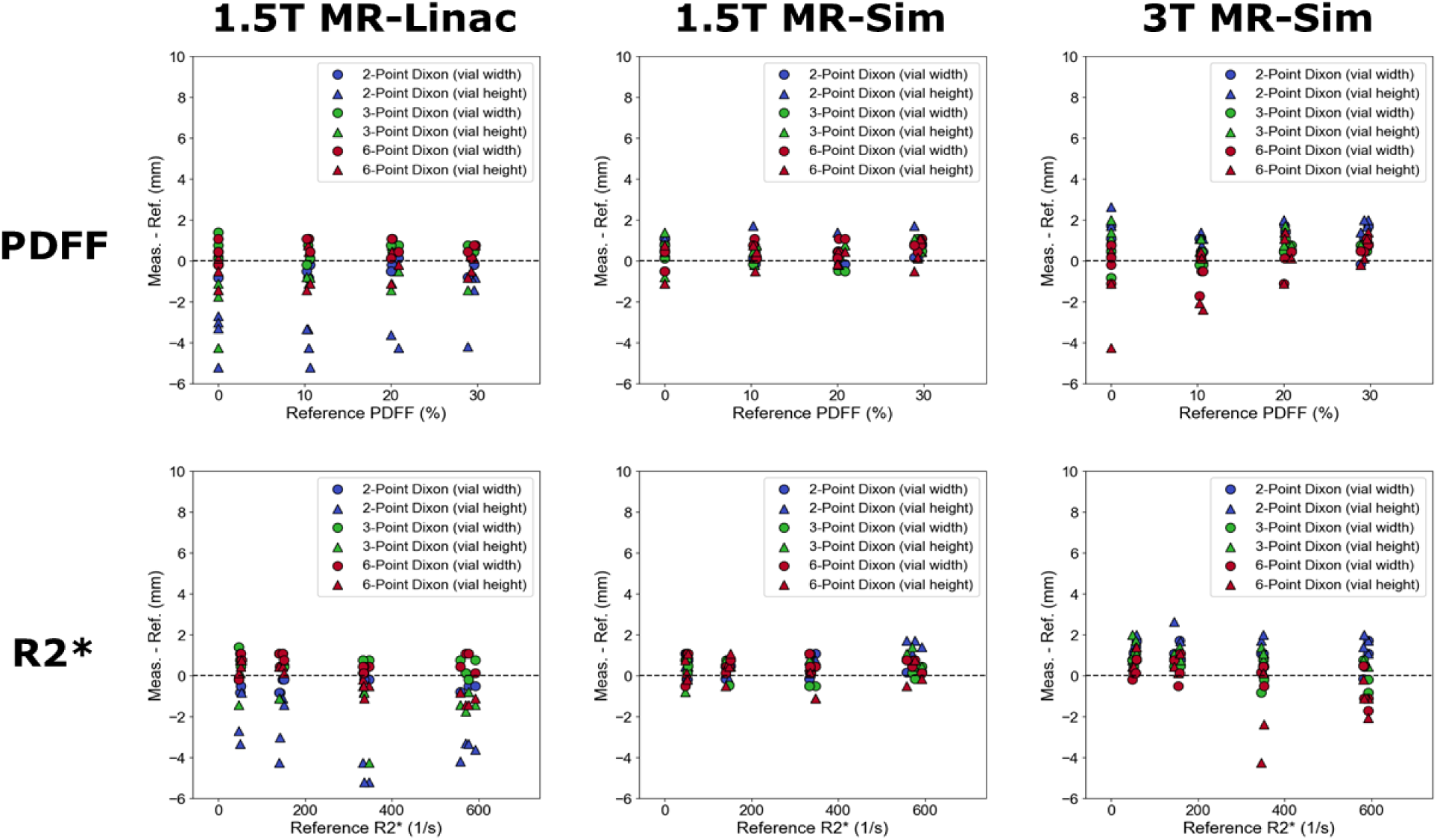
The quantified geometric distortion represented as the difference between the measured width and height of each vial in the Calimetrix phantom and the nominal diameter of 25 mm across each scanner and Dixon sequence tested.

The concordance plots for both the PDFF and R2* across all three scanners tested in this study is shown in **Figure 4**. For the 2-point Dixon sequence, the PDFF values are overestimated across all reference values on the 1.5T MR-Linac while showing mixed positive and negative bias on both MR-Sim systems. The 3-point Dixon sequence showed similar overestimation of PDFF values on the 1.5T MR-Linac but at a lower magnitude while the 1.5T MR-Sim was slightly overestimated at lower PDFF values and underestimated at higher PDFF values. The 3T MR-Sim did not show any visible PDFF bias. Across all scanners tested, the 6-point Dixon sequence showed the least PDFF bias. For R2*, both the 3-point and 6-point Dixon sequences showed similar mean bias, however the 6-point sequence seemed to be more consistent across the five repetitions for all scanners tested.

**Figure 4.**
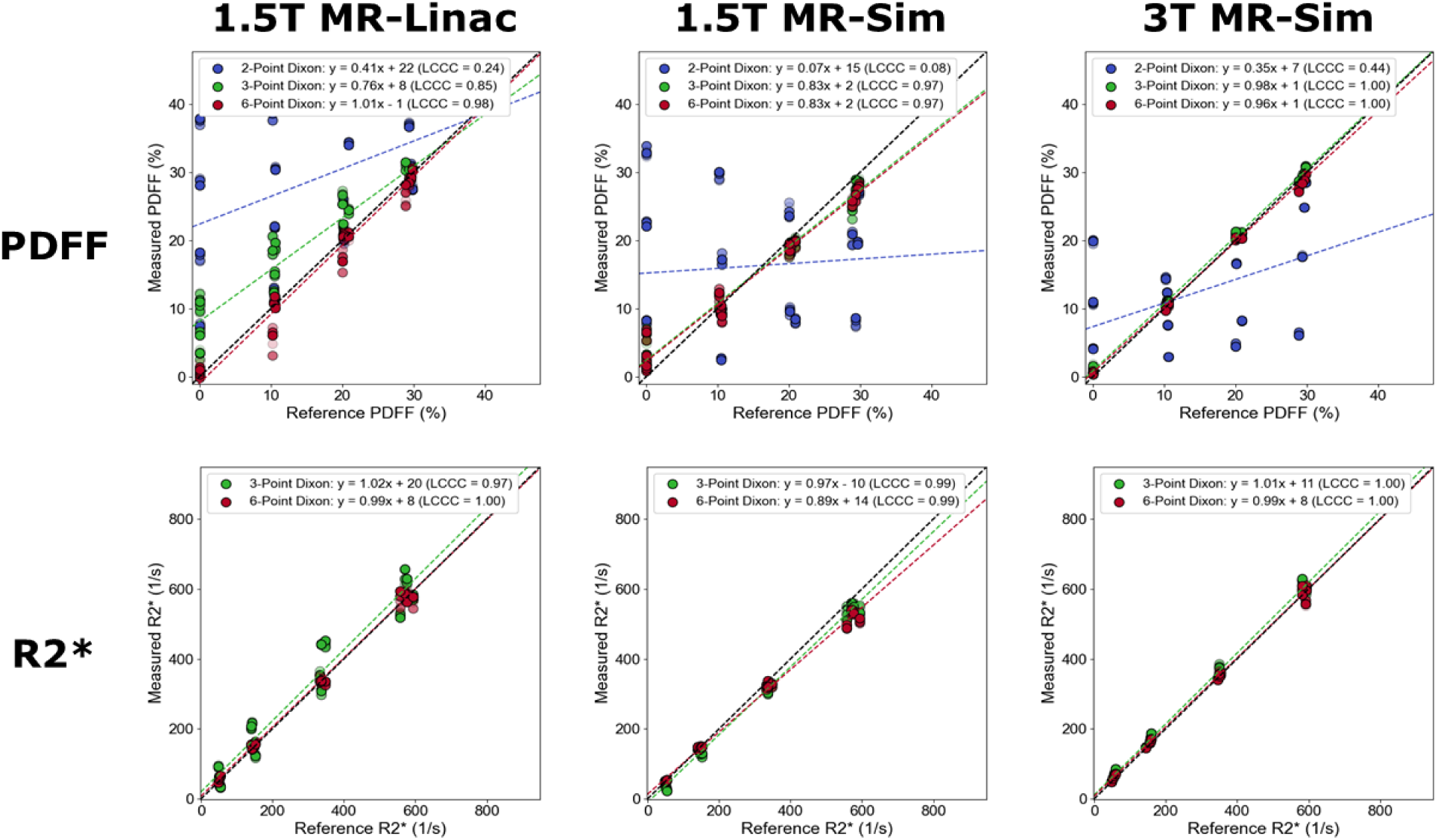
Concordance plots of the quantitative R2* and PDFF values from the proposed quantitative Dixon sequences compared to the phantom nominal reference values. The opacity of the scatter dots is used to separate each of the five repetitions. For each series of data, a linear line of best fit (solid black line) was computed along with its 95% confidence intervals (dashed red lines).

The results of the Bland-Altman analysis for both PDFF and R2* for the 2-/3-/6-point Dixon sequences across all scanners tested are shown in **Figure 5**. The 2-point Dixon sequence showed the highest variability from phantom reference values for PDFF. Quantitatively, the 95% limits of agreement for the 2-point Dixon sequence had a range of 22, 27, and 20% for the 1.5T MR-Linac, 1.5T MR-Sim, and 3T MR-Sim, respectively. Comparatively, the 3-/6-point Dixon sequences had a range of 7/5%, 4/4%, and 1/2%, respectively. For the R2* Bland-Altman analysis, the 95% limits of agreement for the 3-/6-point Dixon sequences had a range of 88/25 1/s, 29/51 1/s, and 25/23 1/s, respectively. All limits of agreement for both PDFF and R2* include 0%.

**Figure 5.**
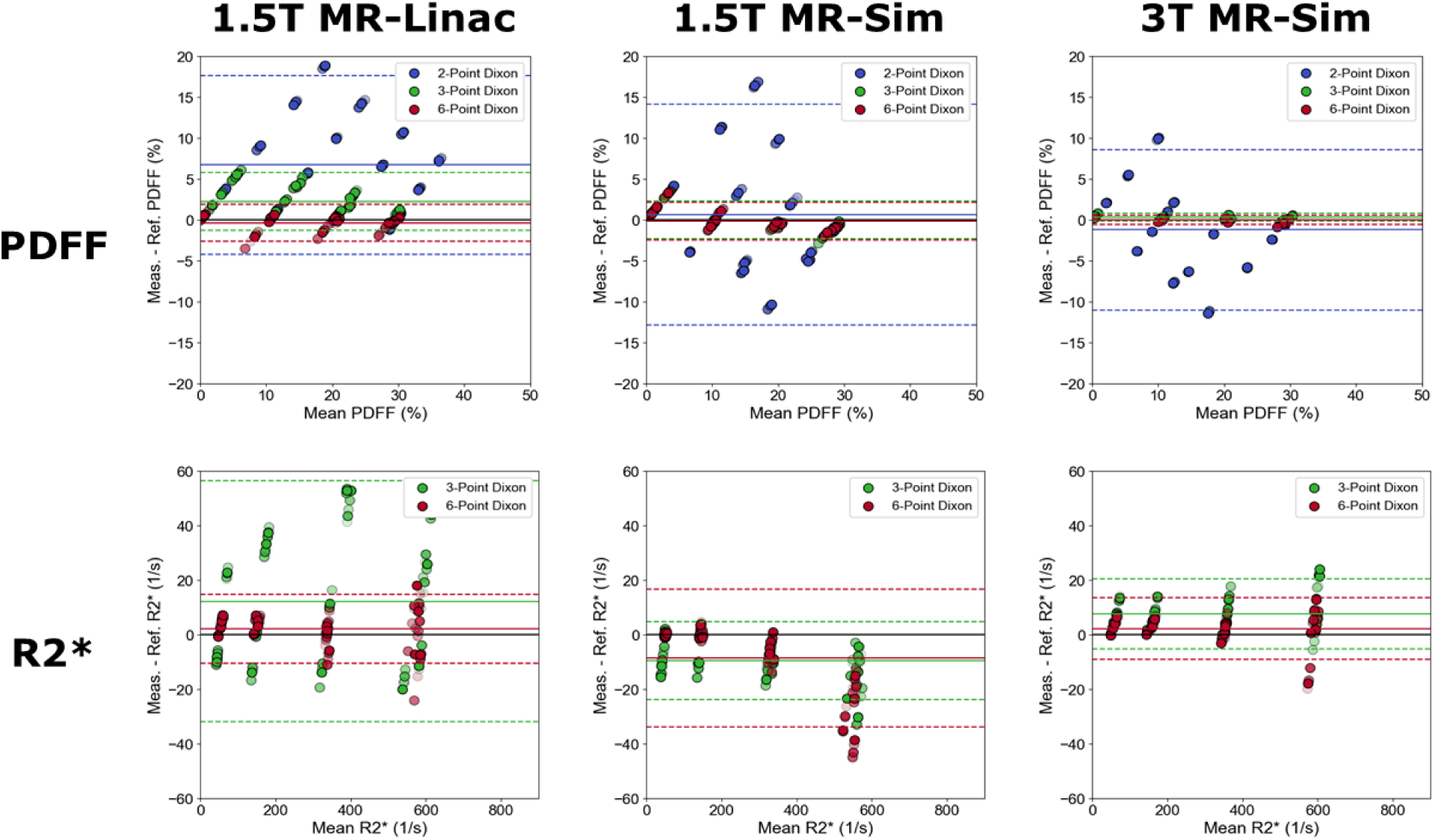
Bland-Altman analysis for both PDFF and R2* for the 2-/3-/6-point Dixon sequences across each scanner tested. The opacity of the scatter dots is used to separate each of the five repetitions. For each plot and sequence type, the mean (solid line) was computed along with its 95% limits of agreement (dashed lines).

A visualization of the PDFF and R2* bias across reference PDFF and R2* combinations in shown in **Figure 6**. Across all scanners, the lower R2* values led to the lowest PDFF bias. On the 1.5T MR-Linac, the higher PDFF values regardless of the R2* value led to the lowest PDFF while on the 1.5T and 3T MR-Sim scanners, increasing values of R2* led to monotonically increasing bias in PDFF. For R2*, generally the lower PDFF values led to lower R2* bias with the highest bias being seen at the combination of high PDFF and high R2*.

**Figure 6.**
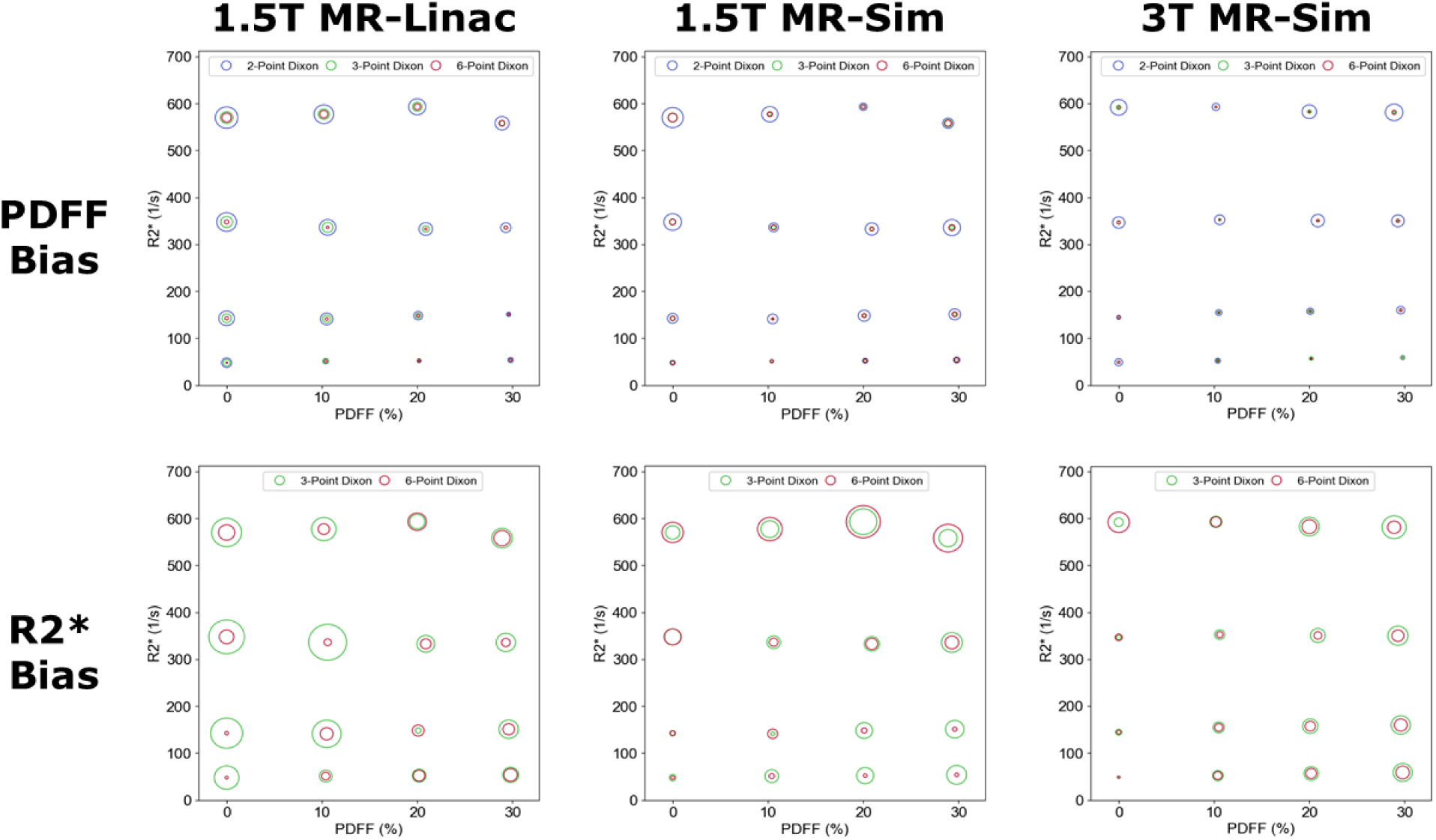
The measured bias against the phantom reference values across each scanner and Dixon sequence tested where the larger circles represent a larger absolute bias. For PDFF the low R2* values lead to the lowest level of PDFF bias while the combined low R2* and low PDFF lead to the lowest level of R2* bias.

The goodness-of-fit plots for the 1.5T and 3T MR-Sim are shown in **Figure 7**. It should be noted that a lower goodness-of-fit is desired since this is a representation of the residual fitting error. Generally, the 3T MR-Sim had lower goodness-of-fit than the 1.5T MR-Sim while the 6-point Dixon was higher than the 3-point Dixon. More specifically, the goodness-of-fit was lowest at higher PDFF values and lower R2* values for both the 3-point and 6-point Dixon sequences, however this difference was more sensitive and prominent with the 3-point Dixon sequence.

**Figure 7.**
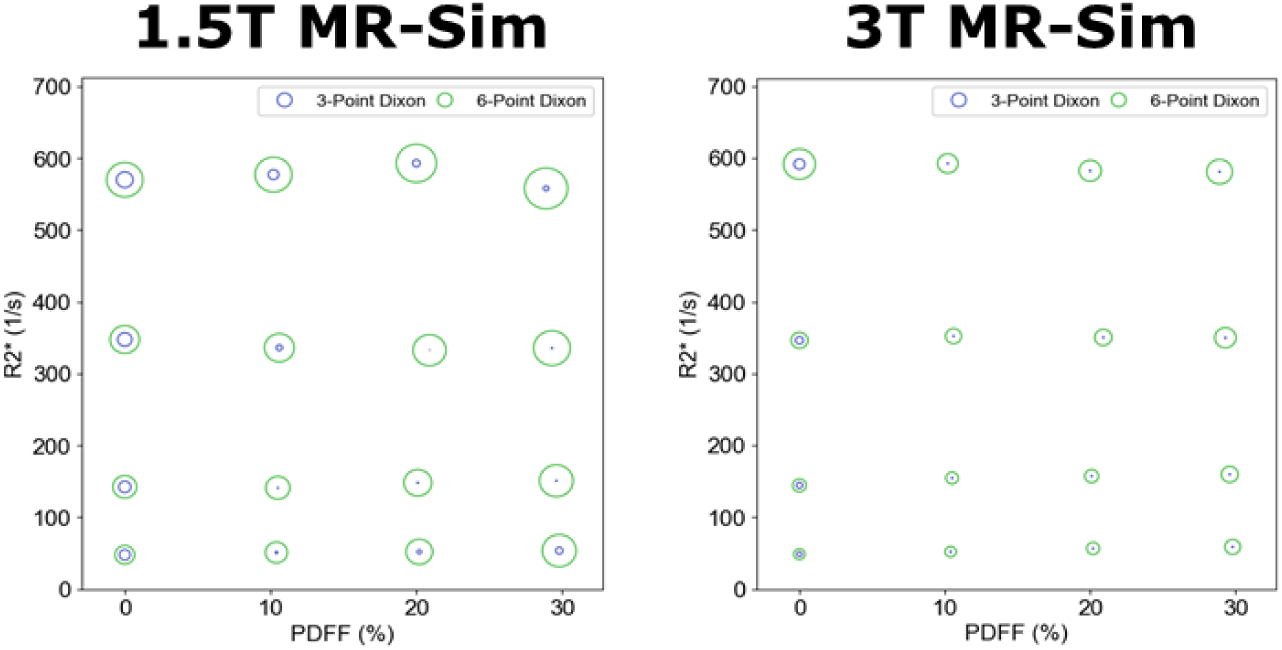
The goodness-of-fit plots for the 1.5T and 3T MR-Sim scanners. Note, the 1.5T MR-Linac did not have a goodness-of-fit automatic reconstruction available from the scanner.

The repeatability CoV of each Dixon acquisition across all three scanners tested is shown in **Figure 8**. For both PDFF and R2*, the 6-point Dixon acquisition consistently has the lowest repeatability coefficient-of-variation (CoV) except for the 1.5T MR-Linac since its measured PDFF was closer to the reference values of near 0%. For each of the three acquisitions, the higher PDFF leads to lower repeatability CoV while R2* stayed similar across the range of R2* values. However, for both the 1.5T MR-Linac and 1.5T MR-Sim, the R2* repeatability CoV was higher at the lowest R2* value.

**Figure 8.**
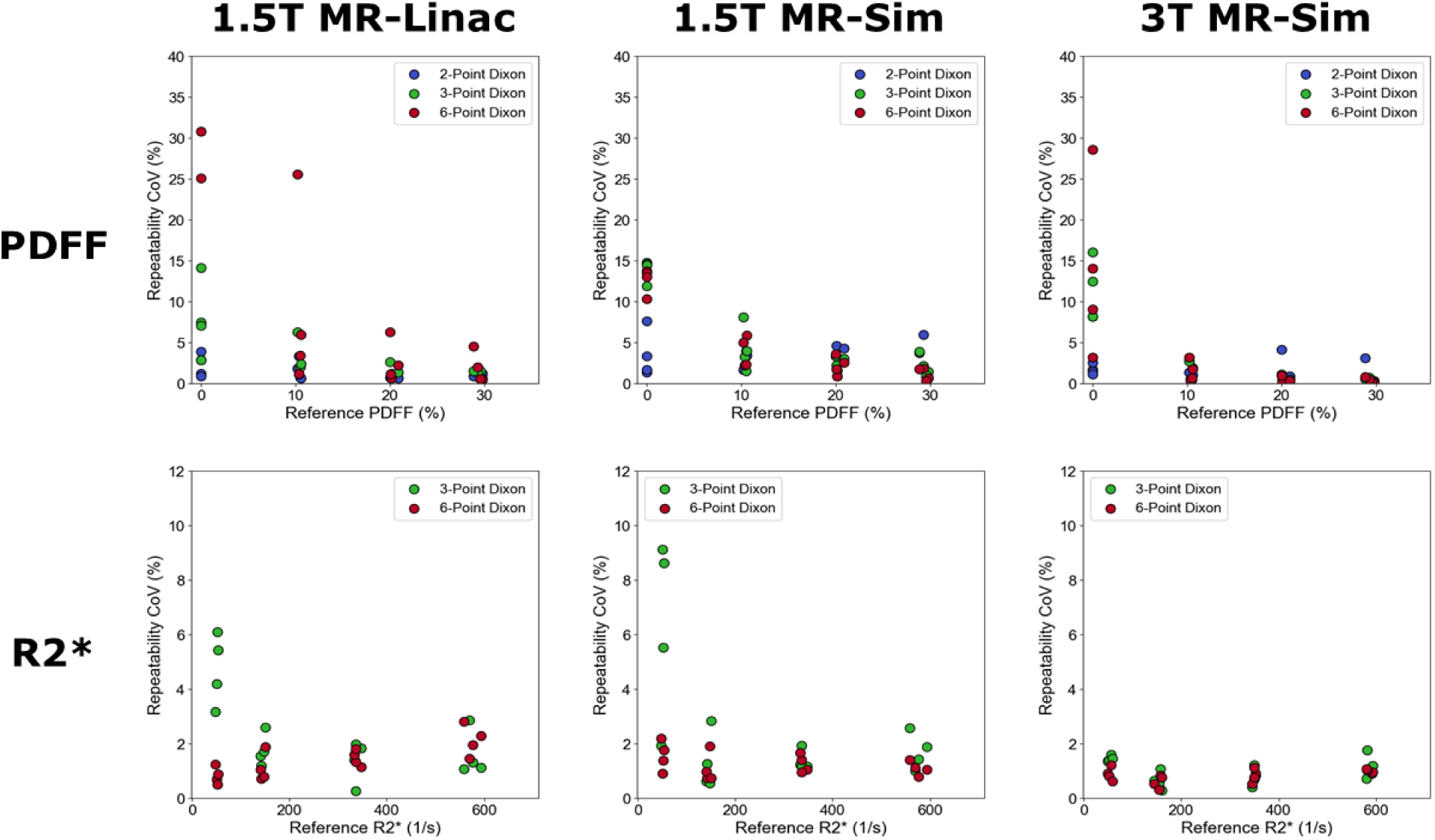
Repeatability analysis across each scanner tested for each of the phantom reference values averaged across all five repetitions.

The reproducibility CoV of each Dixon acquisition across all three scanners tested is shown in **Figure 9**. General trends are that the 6-point Dixon sequence had a lower reproducibility CoV than the 3-point Dixon sequence which was lower than the 2-point across both PDFF and R2*. The magnitude of the reproducibility CoV generally decreased across both higher PDFF and R2* due to the higher percentage changes possible at low reference values.

**Figure 9.**
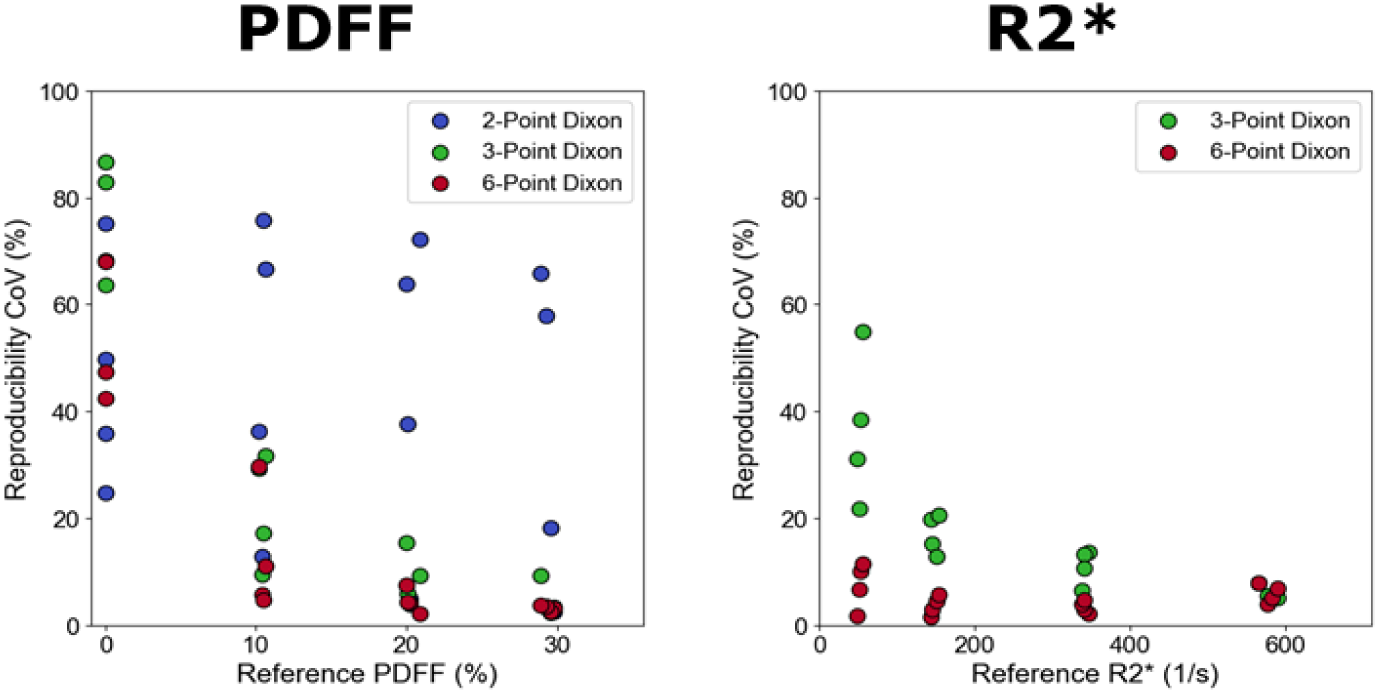
Reproducibility analysis across each scanner tested for each of the phantom reference values averaged across all five repetitions. For both PDFF and R2*, the 6-point Dixon acquisition consistently has the lowest reproducibility coefficient-of-variation (CoV). For each of the three acquisitions, the higher PDFF and R2* leads to lower reproducibility CoV.

A comparison between the measured PDFF and R2* values between the 3T MR-Sim and 1.5T MR-Sim, 1.5T MR-Sim and 1.5T MR-Linac, and 3T MR-Sim and 1.5T MR-Linac is shown in **Figure 10**. When comparing the 6-point Dixon sequence on the 3T MR-Sim to the 1.5T MR-Sim, both the PDFF and R2* were higher in the 3T MR-Sim with a slope of 1.13 and 1.12, respectively. This trend continued for the 3-point Dixon sequence. Second, when comparing the 6-point Dixon sequence on the 1.5T MR-Sim to the 1.5T MR-Linac, both the PDFF and R2* were lower in the 1.5T MR-Sim with a slope of 0.78 and 0.90, respectively, while for the 3-point Dixon sequence the PDFF was slightly higher on the 1.5T MR-Sim with a slope of 1.03 while the R2* slope was still 0.92. Finally, when comparing the 6-point Dixon sequence on the 3T MR-Sim to the 1.5T MR-Linac, the PDFF was lower in the 3T MR-Sim while the R2* was slightly higher with a slope of 0.92 and 1.01, respectively. This trend was reversed in the 3-point Dixon sequence with a slope of 1.20 and 0.96, respectively.

**Figure 10.**
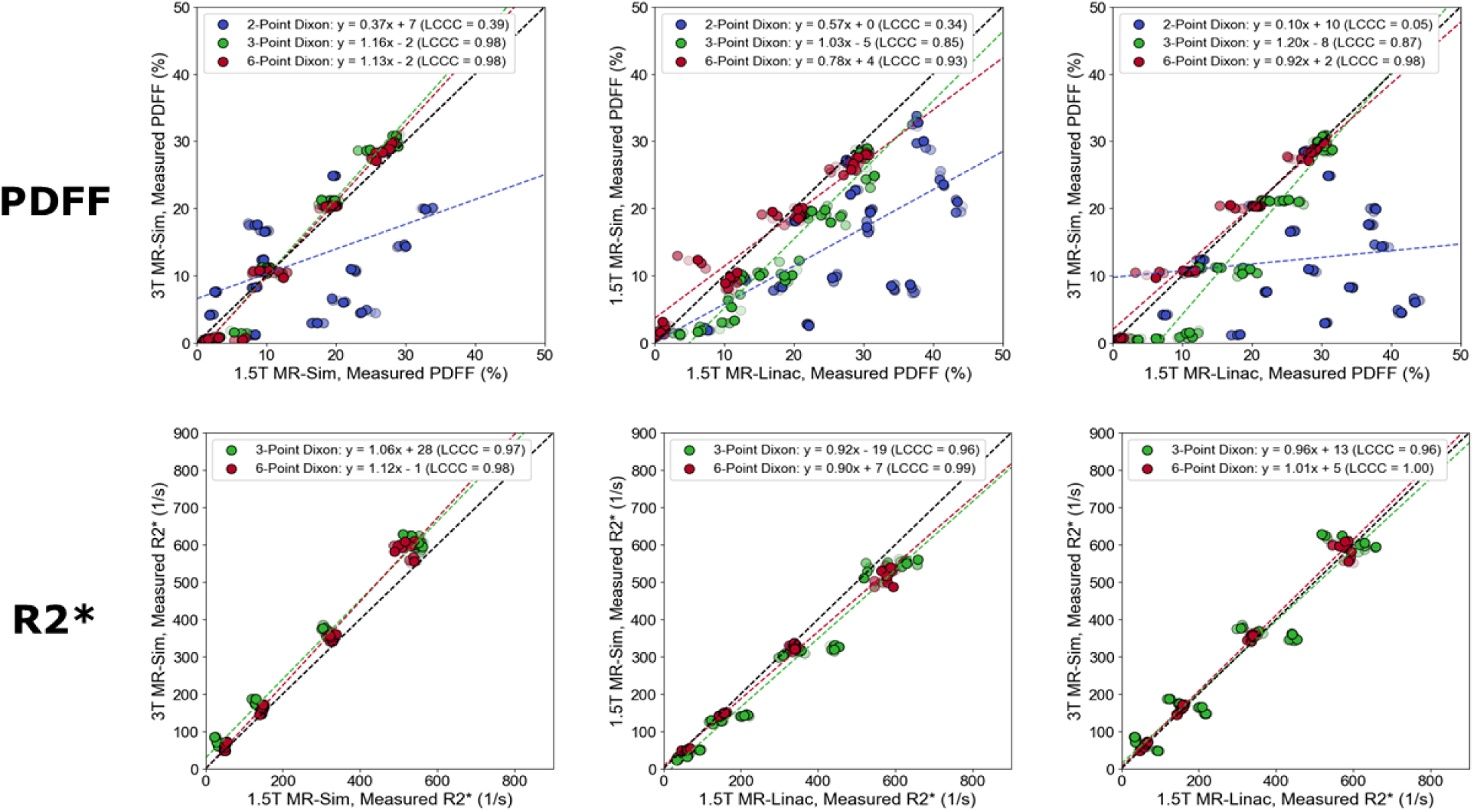
A comparison between the measured PDFF and R2* values on the 3T MR-Sim and 1.5T MR-Sim (left), 1.5T MR-Sim and 1.5T MR-Linac (middle), and 3T MR-Sim and 1.5T MR-Linac (right).

### 3.3. Healthy Volunteer / Patient Results

A qualitative comparison of the change in PDFF and R2* quantitative maps across the 2-/3-/6-point Dixon sequences in a healthy volunteer on the 1.5T MR-Linac is shown in **Figure 11**. Visually, the PDFF values in the muscle appear to be overestimated in the 2-point Dixon sequence with low expected PDFF while being underestimated in high expected PDFF compared to the 6-point Dixon sequence. Quantitatively, the mean [standard deviation] PDFF in: 1) iliacus muscle is 16 [6], 4 [10], and 2 [0]%, 2) yellow bone marrow is 82 [6], 92 [9], and 92 [6]%, 3) red bone marrow is 66 [8], 65 [9], and 68 [8]%, and 4) subcutaneous adipose tissue is 83 [7], 85 [8], and 90 [7]% for the 2-/3-/6-point Dixon sequences, respectively. For the R2* quantitative maps, the 3-point Dixon sequence showed a prominent decrease in spatial resolution and contrast compared to the 6-point sequence. Quantitatively, the mean [standard deviation] R2* in: 1) iliacus muscle is 95 [10] and 37 [19] 1/s, 2) yellow bone marrow is 99 [8] and 38 [27] 1/s, 3) red bone marrow is 117 [17] and 78 [60] 1/s, and 4) subcutaneous adipose tissue is 135 [41] and 18 [16] 1/s for the 3-/6-point Dixon sequences, respectively.

**Figure 11.**
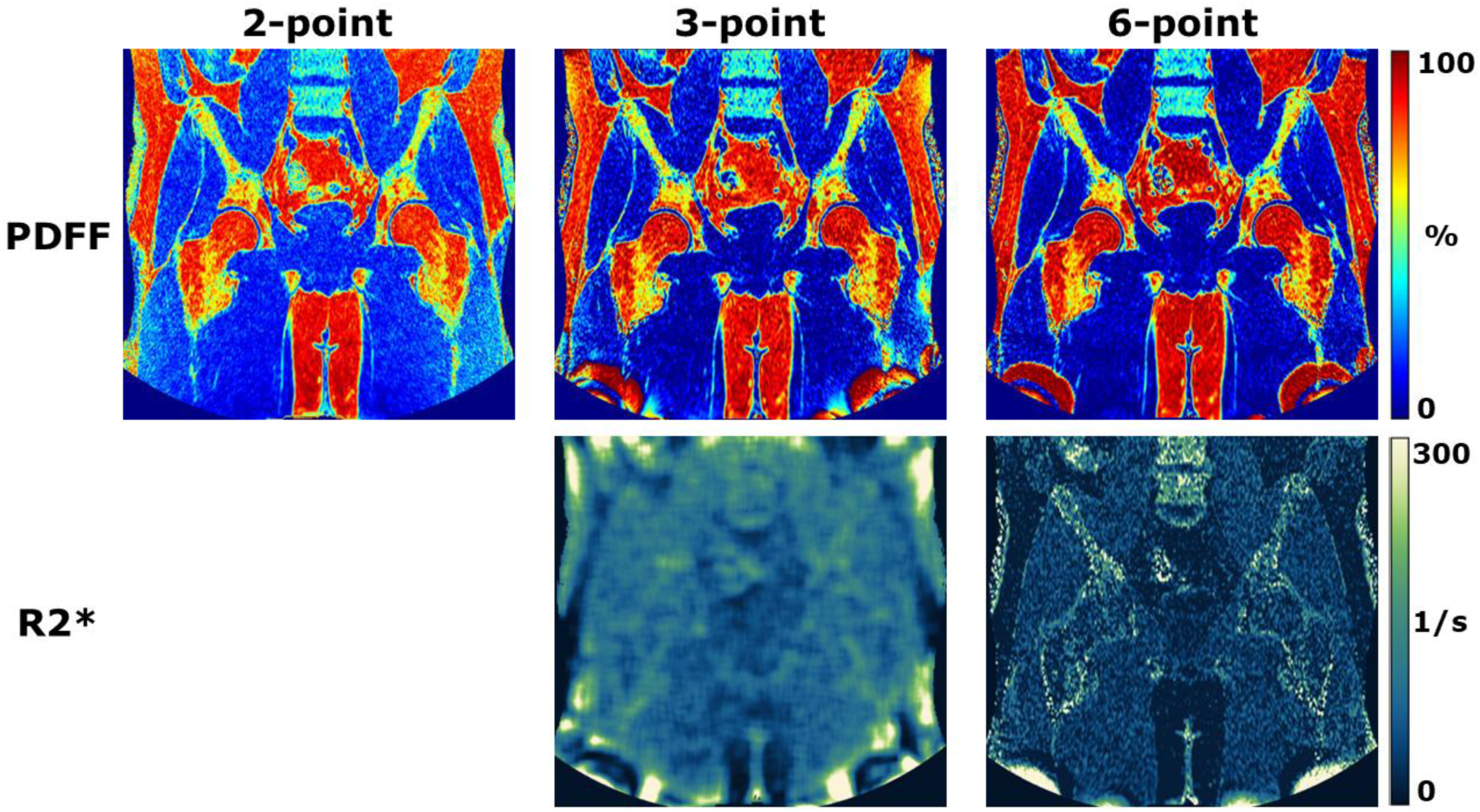
A demonstration of the change in PDFF and R2* values across the 2-/3-/6-point Dixon sequences in a healthy volunteer on the 1.5T MR-Linac. The R2* maps from the 3-point Dixon sequence showed degraded spatial resolution and contrast compared to the 6-point Dixon sequence.

The optimal 6-point Dixon sequence across one healthy volunteer (1.5T MR-Linac) and two prostate cancer patients (1.5T/3T MR-Sim) for all scanners tested is shown in **Figure 12**. Visible red bone marrow is seen for each patient, identified as the regions inside the bone with the lower PDFF values. Interestingly, the 3T MR-Sim prostate cancer patient shows almost complete L5 yellow bone marrow while L4 is almost complete red bone marrow while for the other scanners, both L4 and L5 show almost complete red bone marrow. The quantitative R2* maps seem to show sometimes orthogonal contrast information when compared to the PDFF maps.

**Figure 12.**
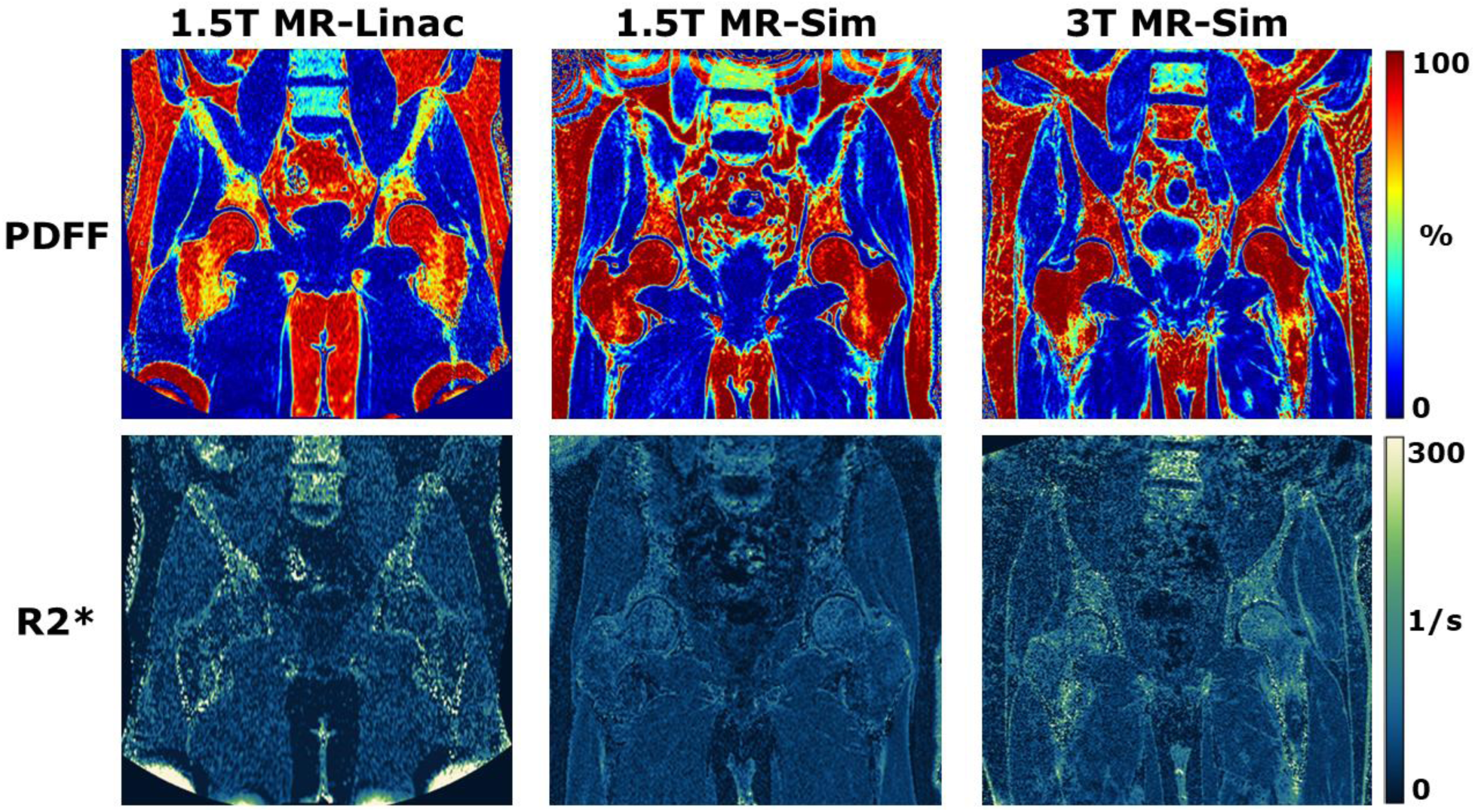
A demonstration of the change in PDFF and R2* values for the 6-point Dixon sequence across one healthy volunteer (1.5T MR-Linac) and two prostate cancer patients (1.5T/3T MR-Sim). A noted variation in the quantity and distribution of red bone marrow can be seen across the quantitative PDFF maps in the femur and pelvis bone.

The optimal 6-point Dixon sequence across one healthy volunteer (1.5T MR-Linac) and two head and neck cancer patients (1.5T/3T MR-Sim) for all scanners tested is shown in **Figure 13**. A visible tumor with high fat content and restricted T2* decay can be seen on the patient’s left anterior side on the 3T MR-Sim scans. In normal tissue areas such as the masseter muscles, parotid glands, and subcutaneous adipose tissue, general agreement in the PDFF values is seen while the R2* maps are more difficult to interpret. The 1.5T MR-Sim R2* map appears to show the highest level of image noise.

**Figure 13.**
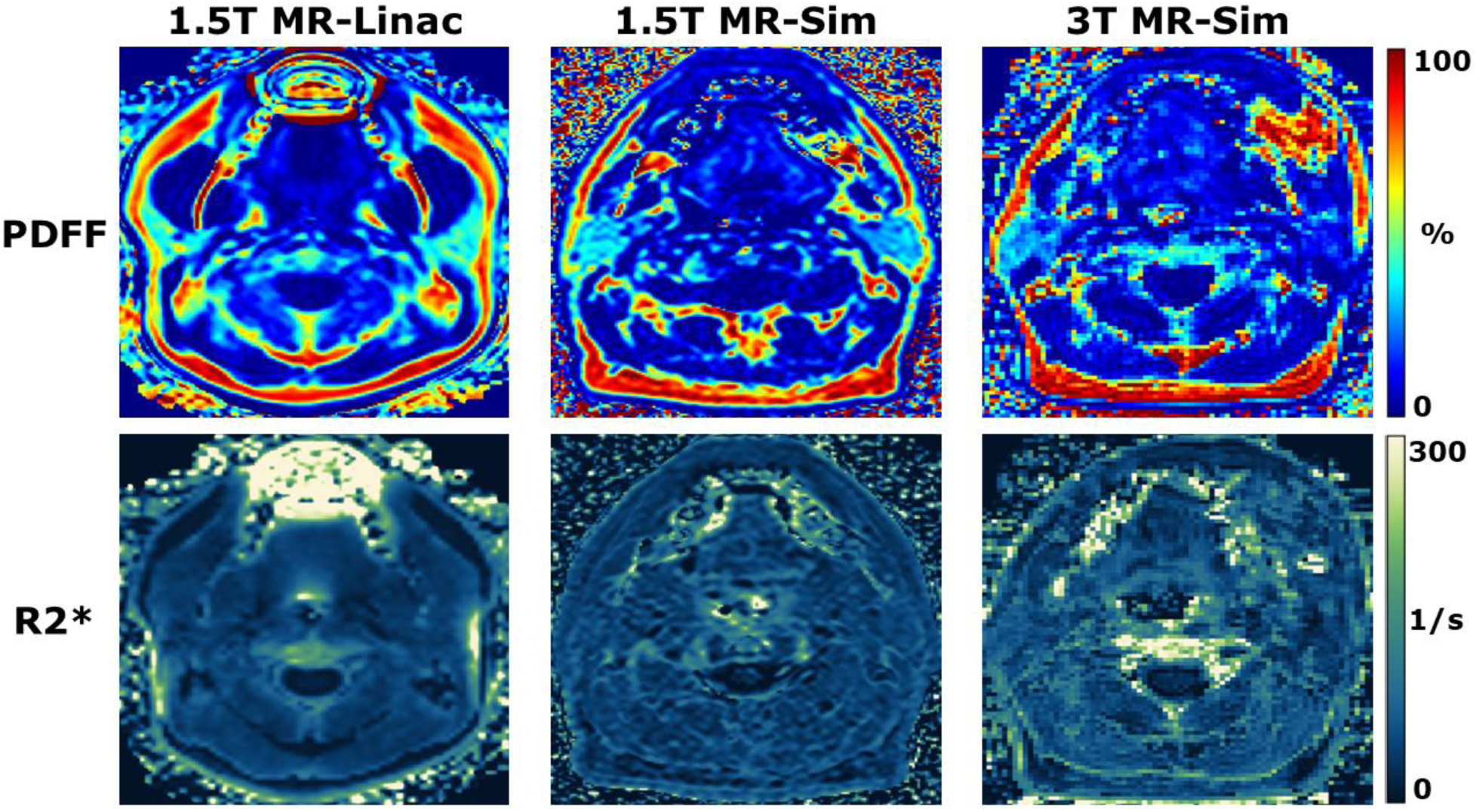
A demonstration of the change in PDFF and R2* values for the 6-point Dixon sequence across one healthy volunteer (1.5T MR-Linac) and two head and neck cancer patients (1.5T/3T MR-Sim).

## 4. Discussion and Conclusion

This work investigated the technical feasibility, repeatability, reproducibility and optimization of simultaneously quantifying R2* and PDFF using 2-point, 3-point, and 6-point quantitative Dixon acquisitions across a 1.5T MR-sim, 3T MR-sim, and 1.5T MR-Linac. This study includes several impactful contributions to the field of MRI-guided radiation therapy including: (1) the first published comparison of 2-point, 3-point, and 6-point Dixon acquisitions on both the MR-sim and MR-Linac scanners, (2) the second^75^ published evaluation of the performance of mDIXON-Quant on the 1.5T MR-Linac, and (3) the first to directly compare it to the rest of the MR-guided radiation therapy fleet (i.e., MR-sim).

The most severe geometric distortion was on the 1.5T MR-Linac when using the 2-point Dixon sequence with distortions of up to 5 mm or more. This could be due to increased complexity of the MR-Linac system which needs to account for the integrated radiation therapy delivery components. This geometric distortion was reduced to under 2 mm for the other tested scanner and specific Dixon sequence combination except in a few outlier scenarios. Specifically, the 3-/6-point Dixon sequences have an enhanced ability to estimate the static magnetic field and correct for any susceptibility-induced geometric distortion^76,77^. For the concordance analysis, the 6-point Dixon sequence showed the highest LCCC at above 0.97 across all scanners for both PDFF and R2*. The 3-point Dixon sequence showed similar average linearity; however, its concordance was slightly lower due to the wider range in values across repetitions compared to the 6-point Dixon sequence. The 2-point Dixon sequence exhibited significant PDFF biases particularly at the higher R2* values since it did not correct for it during reconstruction. For the Bland-Altman analysis, the 2-point Dixon sequence had the widest 95% limits of agreement with the 3-point being next followed by the 6-point Dixon sequence with the narrowest bands. A notable exception was seen in the 1.5T MR-Sim where the R2* 95% limits of agreement were wider in the 6-point Dixon sequence compared to the 3-point Dixon sequence. All 95% limits of agreement for each Dixon sequence on each scanner for both PDFF and R2* included the 0-value indicating no significant level of systematic bias. Although the calculated goodness-of-fit was higher in the 6-point Dixon compared to the 3-point Dixon, this could be due to the higher number of echoes which increases the potential for fitting error, especially in the case of hitting the noise floor at the higher echo times^78^. The primary finding was that the goodness-of-fit is generally lowest at higher PDFF values and lower R2* values. This variability was more pronounced in the 3-point Dixon sequence, indicating its instability across a wide range of clinically relevant PDFF and R2* combinations. In general, the repeatability CoV was the lowest for the 6-point Dixon sequence across both PDFF and R2*; however, it showed extremely high values on the 1.5T MR-Linac for PDFF since the measured values were low and small deviations lead to large calculated CoV. The 6-point Dixon sequence showed the lowest reproducibility CoV averaging under 10% compared to the 3-point Dixon sequence which was nearly twice as high across both PDFF and R2* values. The slope when comparing the PDFF and R2* values from the 6-point Dixon sequence across scanners was as low as 0.78 (1.5T MR-Sim vs. 1.5T MR-Linac PDFF) and as high as 1.13 (3T MR-Sim vs. 1.5T MR-Sim PDFF) indicating the potential for bias when comparing the same patient across different scanners. In addition to these agreements with published reference values, the 6-point Dixon sequence also has double the NSA compared to the 3-point Dixon sequence and for a wider range of dephasing angles which further motivates its clinical use in place of both the 2-point and 3-point Dixon sequences.

In the pelvis, the yellow bone marrow PDFF was around 92% for the 6-point Dixon sequence while monotonically decreasing to 82% for the 2-point Dixon sequence while the red bone marrow was around 66% across all Dixon sequences. This agreed with published reference values which typically places a 70% PDFF cutoff threshold for determination of red bone marrow^24^. We found R2* values in the red bone marrow of 78 1/s using the 6-point Dixon sequence while reported literature^79,80^ has shown around 130 1/s. For our study, the 1.5T MR-Linac healthy pelvis volunteer was younger than the prostate cancer patients which is why more red bone marrow is visible^81^, particularly in the pelvic bone. Typically, a 70% PDFF cutoff is used to separate red bone marrow from yellow bone marrow^24^.

In the head and neck, we found that the quantitative measurements from the PDFF and R2* maps for the 6-point Dixon sequence agreed with published reference values. The parotid PDFF was around 40% whereas literature^82–84^ shows it to be a median around 35% with a wide interquartile range of around 20% while the parotid R2* was around 100 1/s whereas literature^85^ shows it to be around 72 1/s. Our measured masseter muscle PDFF was around 5% whereas literature^84^ shows it to be around 0% and our measured subcutaneous adipose tissue PDFF in both the was around 90% whereas literature^83,84^ shows it to be around 90% as well. Second, the head and neck patient presented here may have reduced PDFF in their left parotid gland due to its proximity to the tumor and previous therapies or possible parotitis^86^.

Native vendor reconstruction was utilized for computing the quantitative R2* and PDFF maps, however a more advanced reconstruction such as Phase Regularized Estimation using Smoothing and Constrained Optimization (PRESCO^87^) may provide more accurate and reproducible results. Further reconstruction methods should be investigated as well and chosen based on optimal clinical workflow and MRI acquisition requirements^88^. More optimizations could be done, such as varying the flip angle to balance signal-to-noise ratio (SNR) with bias introduced from T1 relaxation, varying the receiver bandwidth to optimize for SNR within constraints of the needed echo times, and adjusting the acquired and reconstructed voxel size to balance SNR with spatial resolution. This work should be done in future studies and clinical translations to match the desired clinical need in terms of desired level of anatomical detail and scan time restrictions. The echo times were chosen to optimize for NSA, particularly for the PDFF maps, so further investigation into optimizing these values with optimal R2* mapping within a desired clinical range should be further studied^89^. The most likely reason why the dephasing angles for the 2-point method were substantially lower than the ideal 180° dephasing angle would be due to the use of the Partially-Opposed-Phase (POP^90^) technique which can be seen in the Philips source code and typically aims for 135° dephasing angles to acquire more phase information. At 3T, the required echo times for optimal NSA are reduced by half which was not possible on our systems even with the highest acquisition bandwidth for our desired field-of-view. The echo times could be shifted an entire cycle to achieve the desired peak NSA; however, this would severely limit the R2* quantification accuracy at higher R2* values.

It should be noted that the results presented here are from a small cohort and future work should focus on extending these findings to larger patient cohorts. This specific study should fall into an R-IDEAL Stage 2b or higher methodology to best fit within the requirements for radiation oncology and image-guided cancer treatment in general. Future extensions of this work at the technical level should focus on more advanced MRI-based spectral fat peak decomposition and characterization^91^ to extract the number of double bonds^92^, number of methylene interrupted double bonds, chain length, saturated fatty acids^93^, monounsaturated fatty acids, polyunsaturated fatty acids^94,95^, and ω-3 fatty acids^96^. This in-depth level of quantification allowed by MRI has been shown to be technically, and even clinically feasible in several anatomies critical in radiation oncology such as the breast^97,98^, liver^99–101^, bone marrow^102^, and cardiac tissue^103^. These techniques can be applied to visualize the tumor microenvironment^104^ and unlock an understanding into the critical role of lipids in the development and survival of cancer inside the human body^105^, especially in response to external stressors such as radiation therapy. Further, the integration of quantitative lipid characterization from MRI has been shown to be promising in several studies^106,107^, and should play an important role in the future development of MRI-based quantitative imaging biomarkers, particularly for adaptive radiation therapy. Finally, R2* mapping can play a complimentary role in this by monitoring tumor hypoxia status^108^ throughout radiation therapy, even providing opportunities for advanced control of radiation dose levels given sufficient spatial resolution^109^.

## Contributor Roles Taxonomy (CRediT) Attribution Statement

*Conceptualization*: L.M., N.A.W., and W.F.; *Data curation*: L.M., N.A.W., and W.F.; *Formal analysis*: L.M., N.A.W., K.S., and W.F.; *Funding acquisition*: C.D.F.; *Investigation*: L.M., N.W., K.S., and W.F.; *Methodology*: L.M., N.W., K.S., B.A.T., and W.F.; *Project administration*: A.A., O.S., S.T., J.W., S.S., S.C., C.P.A., C.D.F., and W.F.; *Resources*: B.A.T. and C.D.F.; *Software*: L.M. and N.W.; *Supervision*: A.A., O.S., S.T., J.W., S.S., S.C., C.P.A., C.D.F., and W.F.; *Validation*: L.M. and N.W.; *Visualization*: L.M. and N.W.; *Writing – original draft*: L.M. and N.W.; *Writing - review & editing* L.M., N.A.W., K.S., B.A.T., S.T., A.A., O.S., J.W., S.S., S.C., C.P.A., C.D.F., and W.F.;

## IRB Statement

All participants provided written informed consent. Volunteers were consented to an internal volunteer imaging protocol (PA15-0418), both approved by the institutional review board at The University of Texas MD Anderson Cancer Center.

## Data Availability Statement

All relevant anonymized imaging data and source code necessary for figure reproduction and analysis are available at the following FigShare URL: https://doi.org/10.6084/m9.figshare.31594372 and GitHub repository: https://github.com/Lucas-Mc/quant-dixon_R-IDEAL_0-2a.

## Acknowledgements

None.

## Notes

This manuscript is the result of funding in whole or in part by the National Institutes of Health (NIH). It is subject to the NIH Public Access Policy. Through acceptance of this federal funding, NIH has been given a right to make this manuscript publicly available in PubMed Central upon the Official Date of Publication, as defined by NIH.

